# Meta-analysis on Plasmodium falciparum sulfadoxine-pyrimethamine resistance-conferring mutations in India identifies hot spots for genetic surveillance

**DOI:** 10.1101/2023.07.24.23293131

**Authors:** Abhinav Sinha, Sonalika Kar, Charu Chauhan, CP Yadav, Lokesh Kori

## Abstract

**Background:** India is on track to eliminate malaria by 2030 but emerging resistance to the first-line antimalarials is a recognized threat. As Artesunate+Sulfadoxine-Pyrimethamine (AS+SP) is the drug-of-choice for uncomplicated P. falciparum malaria in most of India, it becomes evident to systematically monitor the validated mutations in Pfdhfr and Pfdhps genes across India. No systematic and robust countrywide surveillance has been reported for these parameters in India.

**Methods:** We included studies that reported data on SP-resistance markers in P. falciparum across India from 2008. Five major databases were exhaustively searched. Individual and pooled prevalence estimates of mutations were obtained through random- and fixed-effect models. The study is registered with PROSPERO (CRD42021236012).

**Results:** A total of 37 publications with data from 80 districts, 21 states and 3 UTs were included. The two PfDHFR mutations, C59R and S108N were the most prevalent mutations and appear to be stabilized/fixed. Although rarest overall, the prevalence of I164L was observed to be as high as 32%. The PfDHFR double mutants were the most prevalent overall. The prevalence of triple and quadruple mutations was 6% and 5%, respectively which is an area of immediate concern. For PfDHPS, the most prevalent mutation was A437G. For PfDHFR/PfDHPS quintuple and sextuple mutations, it was observed that despite a low overall prevalence of these mutations, some areas are critical for surveillance.

**Conclusion:** The analysis brings forward the SP-resistance hot spots and emphasizes critical gaps, and challenges, and suggests focal and local malaria genetic surveillance (including drug resistance markers) till malaria is eliminated.

**Key points:** This manuscript concludes that currently used first-line drugs for P. falciparum malaria, particularly sulfadoxine-pyrimethamine, have a high likelihood of failing in near future. Although currently effective, the evidence is backed by molecular meta-analyses data on drug resistance markers in India. The policy makers and program managers may use this robust data to decide whether it is time to change these currently used anti-malarials in India before it is too late. It also identifies certain hot spots for continuous genetic monitoring of the molecular markers for timely actions.

## 1. Introduction

Oral artemisinin-based combination therapy (ACT) is the treatment of choice for uncomplicated malaria in India: Artemether-Lumefantrine (AL) in the 7 North Eastern Sates (NES) and Artesunate & Sulfadoxine-Pyrimethamine (AS+SP) in the rest of India^1^. The malaria drug policy in India has witnessed two critical changes: switching from chloroquine (CQ) to AS+SP for the entire country in 2010 and then from AS+SP to AL in the NES in 2013.^2, 3^. Both these switchovers were backed by sufficient clinical (therapeutic efficacy studies; TES) and molecular (validated genetic markers for drug resistance) evidence for resistance to CQ and SP.

AS+SP works in tandem: a majority of parasite biomass is quickly eliminated by AS and the residual parasites are killed by the partner drug SP.^4^ The problem arises when either of the two drugs start to fail. In case of artemisinin ‘partial’ resistance leading to delayed parasite clearance, a higher than anticipated parasite biomass is presented to SP that can precipitate ACT failure if SP efficacy is already compromised.^5^ Or, if the partner drug is optimally efficacious, even the presence of artemisinin partial resistance, is not likely to lead to treatment failure^4^, thus signifying the importance of preventing partner drug resistance. On the contrary, even in the presence of optimally efficacious artemisinin derivative, a poorly efficacious partner drug (SP) can lead to ACT failure.^6, 7^ Thus, in order to retain the efficacy of AS+SP, both AS and SP should have optimal efficacies.^8^

Several recommendations to introduce AL across India for uncomplicated Pf-malaria have been made to the Government of India^3^ on the basis of TES and SP-resistance makers prevalence and distribution in India. However, due to a lack of compelling evidence suggesting compromised/declining efficacy of AS+SP, the policy remained unchanged. Although a pre emptive change favouring universal AL across India on the basis of SP-resistance markers may include logistic and financial challenges^3^, the dilemma of whether to switch from AS+SP to AL and when, is worth a reconsideration in the light of accumulating evidence.

It is known that non-synonymous mutations in *Pfdhfr* decrease Pyrimethamine efficacy in a stepwise manner starting with the S108N mutation.^9^ The efficacy further reduces progressively with each of the following substitutions sequentially: C59R, N51I, and I164L^10^ such that the triple (N51I, C59R, S108N) and quadruple (N51I, C59R, S108N, I164L) mutants precipitate SP therapeutic failures. Similarly, A437G alone^11^ or with K540E mutations in *Pf*DHPS are critical for initial development of sulfadoxine resistance^12^ which progresses with additional S436A, A581G and A613S mutations.^13–15^ The *Pf*DHPS mutations appear to get selected in the background of *Pf*DHFR double/triple mutations ^14, 16^ such that the quintuple or the ‘fully resistant genotype’ (N51I-C59R-S108N-A437G-K540E)^17^ and sextuple or the ‘super resistant genotype’ (N51I-C59R-S108N-A437G-K540E-A581G)^17, 18^ mutations confer higher SP resistance than *Pf*DHFR quadruple and independent *Pf*DHPS mutations ^18–20^. The lesser frequent *Pf*DHFR I164L and *Pf*DHPS A581G and A613S appear in the parasite populations once the quintuple *Pf*DHFR+*Pf*DHPS mutants are established in the population and therefore the appearance and rising prevalence of *Pf*DHFR I164L and *Pf*DHPS A581G and A613S mutations alone might be a *sine qua non* of established severe SP resistance.^20, 21^ Therefore, the prevalence and distribution of the *Pfdhfr/Pfdhps* drug resistance markers offer a valid evidence for the underlying SP-resistance and treatment failures.^20, 22–25^ However, their prevalence and spatiotemporal data are scanty, scattered and not uniformly available for India. Only a handful of efforts have been attempted^26–28^ but they suffered from methodological limitations that render them of little utility when it comes to standardization of data for spatiotemporal comparability. This study addresses these limitations and generates robust data synthesis for the program managers to guide evidence-based decisions for a possible shift in India’s malaria drug policy.

## 2. Methods

Search details are shown in Table 1 and Figure 1. Multiple sequence alignment of included NCBI sequences was performed using ClustalW in MEGA X ^29^ considering PF3D7_0417200 and PF3D7_0810800 as references.

**Table 1.** Detailed information showing the search criteria applied to extract information on Plasmodium infection and resistance in India. The first column carries information regarding various search engines and databases used (date-wise). The second and third column shows search term(s) and the field of the article searched, respectively. *Additional search on 15 January 2023 using the same search terms but restricted to years 2020-2023. Multiple sequence alignment of included NCBI sequences was performed using ClustalW in MEGA X^29^ considering PF3D7_0417200 and PF3D7_0810800 as references.

| Search Engines & Databases<br>(date searched) | Search term/s | Field<br>(of the article) searched |
| --- | --- | --- |
| PubMed® Advanced Search<br>Builder (16 August 2020)* | Plasmodium OR Malaria AND<br>Resistance AND India | Title / Abstract |
| Google Scholar Advanced<br>Search (26 December 2020) | Resistance India (with all the<br>words) + Plasmodium<br>malaria (with at least one of<br>the words) | Title |
| Web of Science™ Core<br>Collection Basic Search (2<br>April 2021) | Plasmodium OR Malaria AND<br>Resistance AND India | Topic (Title, Abstract,<br>Keywords) |
| Scopus® Start Exploring (21<br>April 2021) | Plasmodium OR Malaria AND<br>Resistance AND India | Title, Abstract, Keywords |
| Embase® Quick Search (2<br>June 2021) | Plasmodium OR Malaria AND<br>Resistance AND India | Title or Abstract |
| NCBI Nucleotide database (20<br>September, 2020) | dhfr AND Plasmodium AND<br>India; dhps AND Plasmodium<br>AND India | Not applicable |

**Figure 1.**
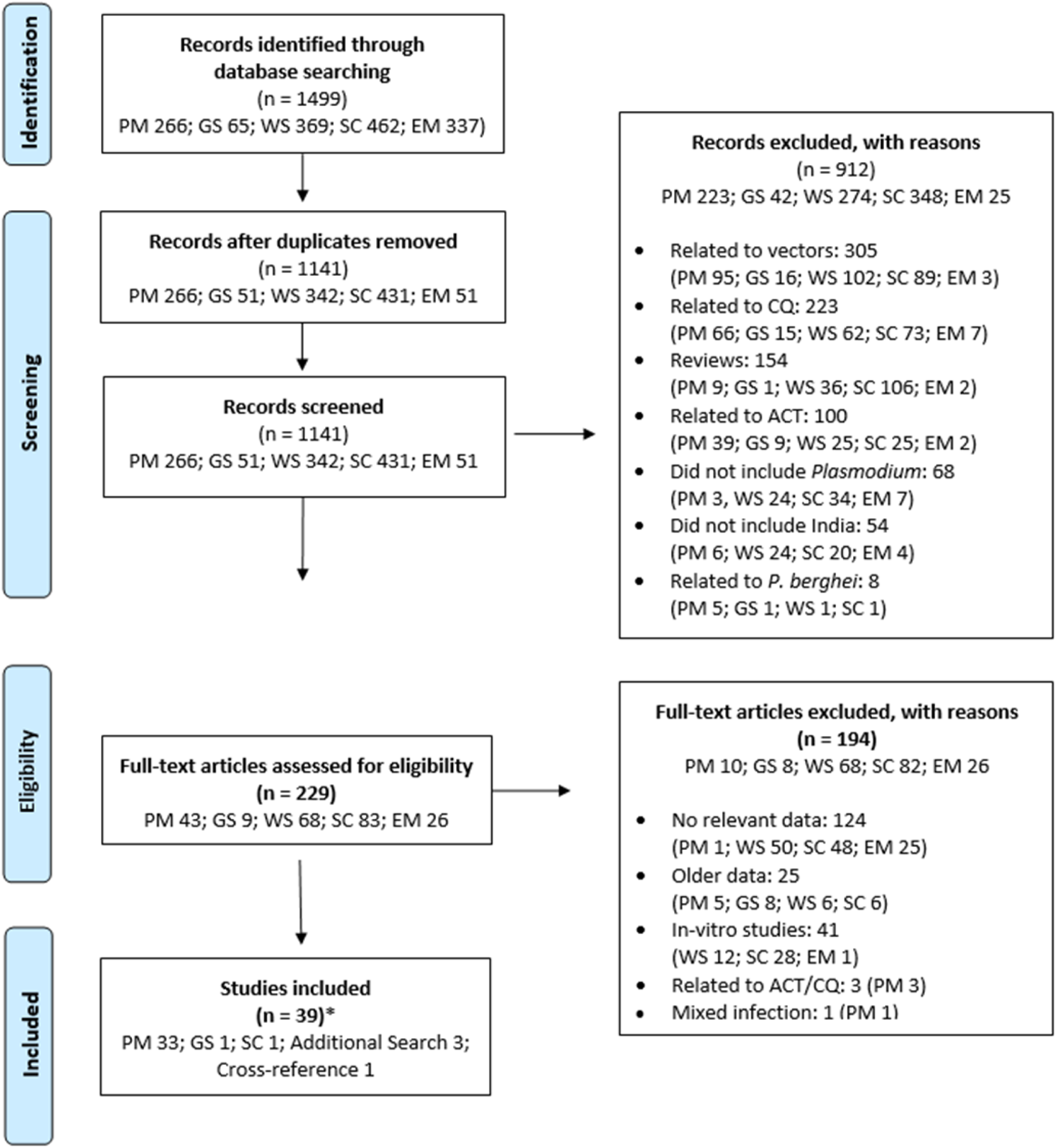
PRISMA (Preferred Reporting Items for Systematic Reviews and Meta-Analyses) flow diagram illustrating the selection of studies for the present systematic review. The study is registered with PROSPERO (CRD42021236012). PM: PubMed® Advanced Search Builder; GS: Google Scholar Advanced Search; WS: Web of Science^TM^ Core Collection Basic Search; SC: Scopus® Start Exploring; EM: Embase® Quick Search; ACT: Artemisinin-based combination therapy; CQ: chloroquine

English-language publications (2008 onwards) and unpublished NCBI database with data on ≥1 *Pf*DHFR/DHPS non-synonymous mutations at desired amino-acid loci were included and pre-2008 data were excluded.

Each included study/sequence was examined independently by AS/LK/SK and CC. Prevalence data were either used as published or extracted and re-analyzed to a uniform format by using the following formula ^18^:

Prevalence of mutation/haplotype = (Number of persons with a particular *Pf* mutation or haplotype / Total number of persons tested for *Pf*)*100

Heterozygous data were considered as mutants. Prevalence up to 1% were considered as 1%.

The following *Pf*DHFR/*Pf*DHPS loci/mutations, validated by WHO ^4, 8, 30^ as SP-resistance markers, were analysed: *Pf*DHFR (N51I/C59R/S108N/I164L); *Pf*DHPS (A437G/K540E/A581G). For analysing the *Pf*DHFR/*Pf*DHPS mutations as markers of degree of SP-resistance, the following criteria were used ^18^: Single mutant (*Pf*DHFR S108N), Double mutants (*Pf*DHFR S108N + *Pf*DHFR N51I OR C59R), Triple mutants (*Pf*DHFR S108N+N51I+C59R), Quadruple mutants (*Pf*DHFR S108N+N51I+C59R+I164L), Quintuple mutants (*Pf*DHFR TRIPLE mutant + *Pf*DHPS A437G+K540E) and Sextuple mutants (QUINTUPLE mutant + *Pf*DHPS A581G). Mutation and haplotype data were included as such or reconstructed to allow comparability.

Data were grouped according to the year of sample collection and name of the states/UTs from where the data was collected. For missing information, the respective supplementary material was explored and corresponding authors were contacted. Data from few of the studies could not be geo-categorized due to non-availability of state-wise breakup and therefore analysed as grouped data. Individual region-wise prevalence data on mutations was chronologically organized from 2008 and presented as percentage.

A geospatial map was prepared using ‘Gramener’ ^31^ that shows the locations (districts) of data collection. Geo-temporal coverage was analysed as temporal depth (TD; number of times a particular State/UT was studied); and spatial depth (SD; number of States/UTs studied in each year). ‘R’ software (version 3.5.2) was used for meta-analyses and creation of forest plots for visualization using “tidyverse”, “meta”, “metafor”. ^32^ Heterogeneity was estimated using the I2 statistic and appropriate modelling (based on I2 value) was done. Both random- and fixed effect models were used as per heterogeneity. Inverse variance random effect model was used for the pooled estimates where I2 was >50%. Individual and pooled prevalence estimates were depicted using forest plots created with a 95 % confidence interval.

## 3. Results

Total 37 publications, 533 *Pfdhfr* and 134 *Pfdhps* NCBI DNA sequences were included (Figure 1). The data (N*Pfdhfr*= 4819-5136 & N*Pfdhps*= 4246-4896 samples, depending upon the type of loci studied) included information from 80 districts, 21 states and 3 UTs (Tables S1a & S2).

The geo-temporal distribution of data (Figure 2a) is not uniform across India and time period included. Temporally, Odisha (OD), West Bengal (WB), Arunachal Pradesh (AR), Chhattisgarh (CG), Tripura (TR), and Assam (AS) were the most frequent (TD score ≥7) locations studied individually whereas for Jharkhand (JK) and Madhya Pradesh (MP), the TD scores were average (4-6). Rest of the regions were studied thrice or lesser which included malaria non endemic zones except the NES of Meghalaya, Mizoram and Nagaland. The combinatorial mutation data for *Pf*DHFR/*Pf*DHPS were available only across 3 time points for JK. Even for the frequently studied states, the data mostly belonged to the year range 2009-2017. Similarly, the years 2009, 2012, and 2014 had comparatively higher SD (number of regions covered).

**Figure 2a.**
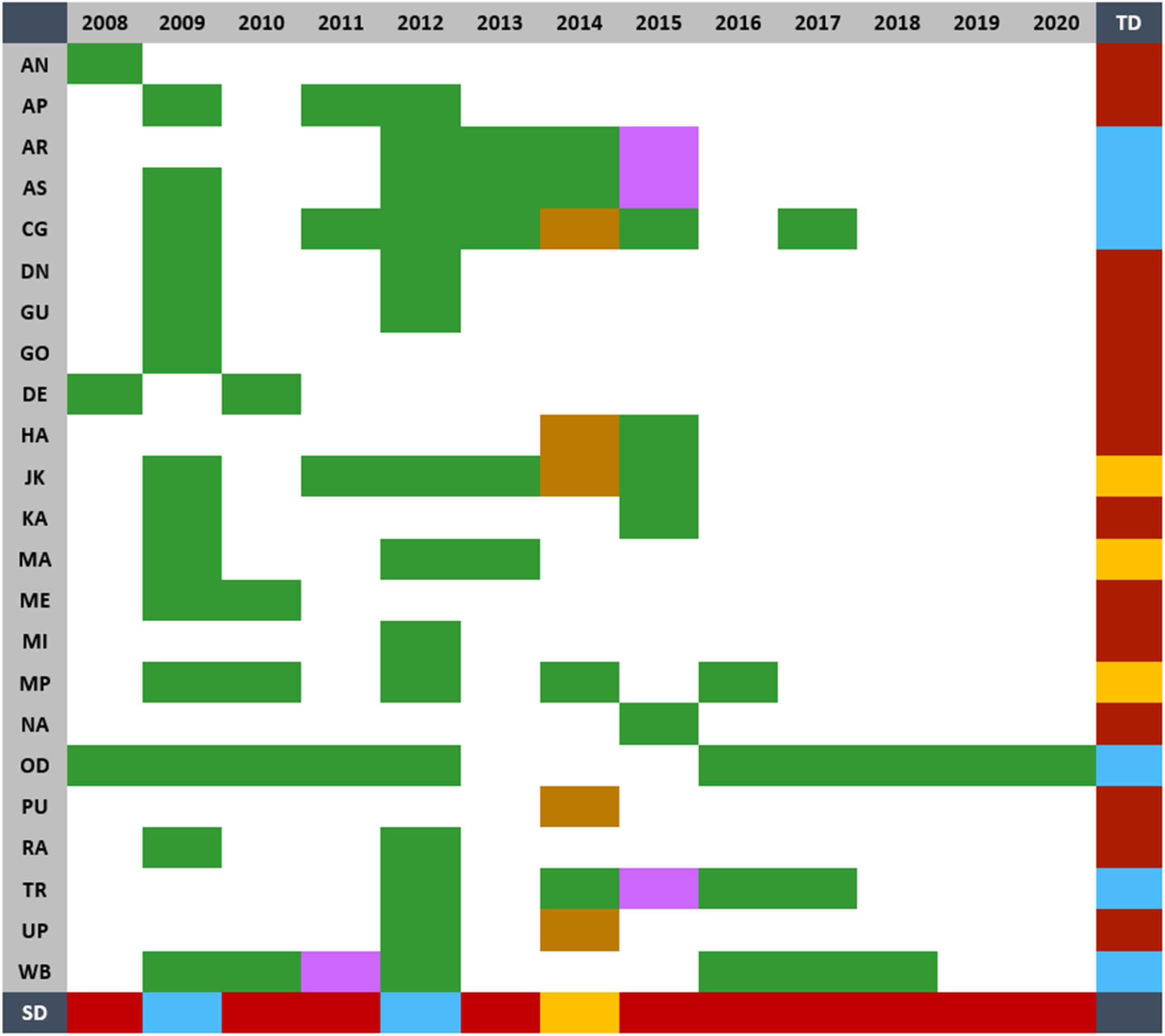
Spatio-temporal depth of data coverage for Pfdhfr and Pfdhps mutations across the studied sites in India. Different States/UTs are arranged in ascending alphabetical order in the first column whereas the years in which they were studied are arranged in ascending order from 2008 to 2020 in the top row. The range of SD was 0 (no State/UT studied) to 23 (all States/UTs studied). For SD, if the State/UT is studied even by >1 researcher in any given year, it was counted as 1. The minimum count for TD was taken as 0 (never studied) with no upper limit. SD scores were classified as poor (0-5), average (6-10) and above-average (11-23) and TD scores were grouped into poor (0-3), average (4-6) and above average (≥7). Green: both Pfdhfr and Pfdhps mutations studied; Pink: only Pfdhps mutations studied; Gold: only Pfdhfr mutations studied; Red: poor score; Yellow: average score; Blue: above-average score. AN: Andaman and Nicobar Islands; AP: Andhra Pradesh; AR: Arunachal Pradesh; AS: Assam; CG: Chhattisgarh; DN: Dadra and Nagar Haveli; GU: Gujarat; GO: Goa; DE: Delhi; HA: Haryana; JK: Jharkhand; KA: Karnataka; MA: Maharashtra; ME: Meghalaya; MI: Mizoram; MP: Madhya Pradesh; NA: Nagaland; OD: Odisha; PU: Punjab; RA: Rajasthan; TR: Tripura; UP: Uttar Pradesh; WB: West Bengal

The geospatial district-wise coverage (Figure 2b) shows that most of the Pf-endemic States/UTs have been covered except the NES (Manipur), however, some states had sparse geo-temporal depth of coverage. Despite being covered in space and time, each state had only few selected districts where such mutations were examined.

**Figure 2b.**
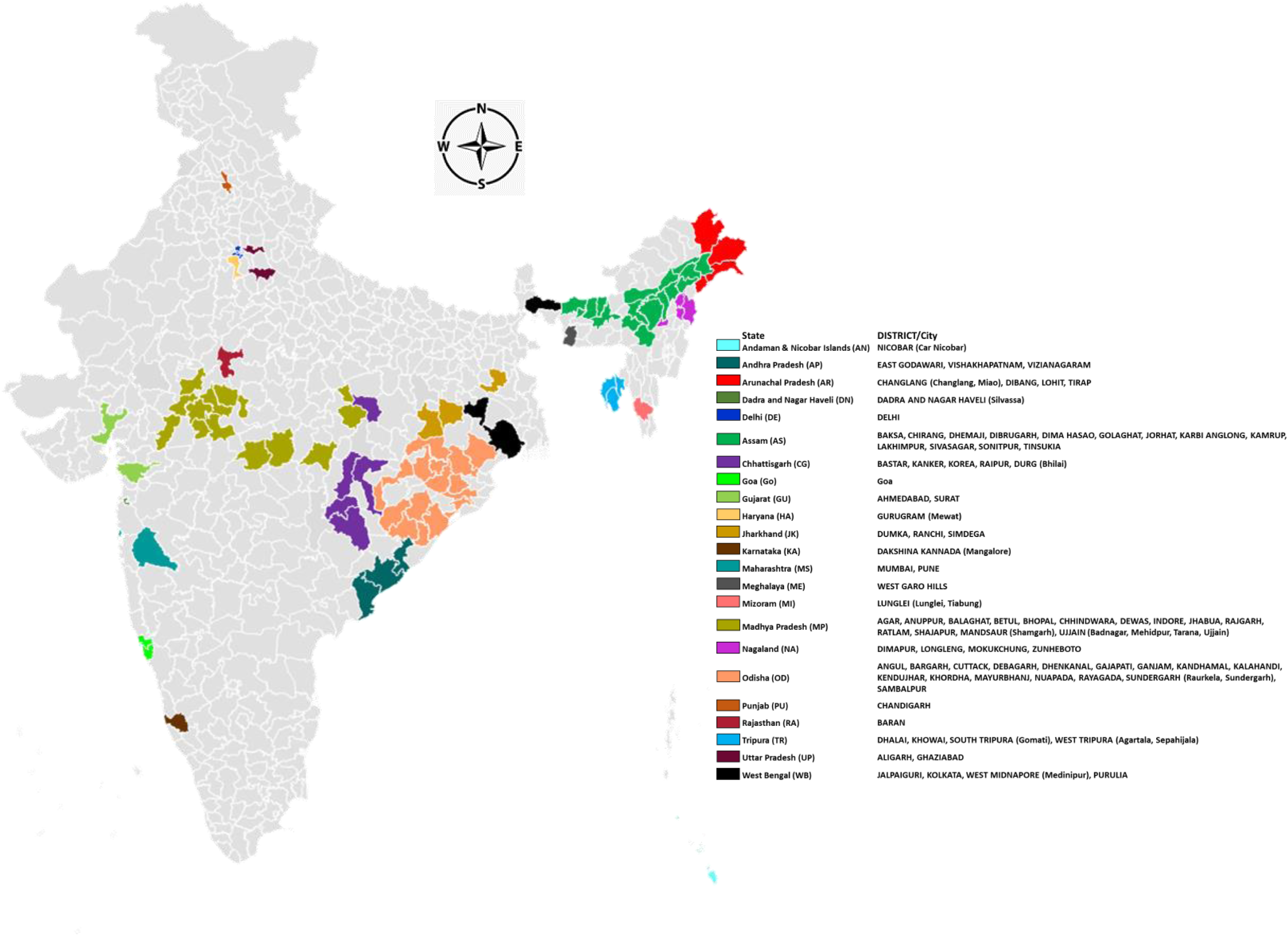
Data collection sites from various districts in India that were included in this study (created with https://gramener.com/map/). The figure depicts data-collection from 80 districts of 23 States and UTs. Different colours represent different administrative units or States/UTs. Different areas of one particular colour represent different districts (sub state administrative units) of that particular state. The colour coding bears the name of the state and their codes followed by the districts (in upper case) and exact site/s of data collection where available (in lower case). **Assam:** In 1995, NORTH CACHAR HILLS district renamed as DIMA HASAO, In 2003, BAKSA was carved out of BARPETA, KAMRUP and NALBARI, In 2004, CHIRANG was carved out of BARPETA, BONGAIGAON and KOKRAJHAR, In 2016, KARBI ANGLONG was divided into WEST KARBI ANGLONG and EAST KARBI ANGLONG; **Jharkhand:** In 2001, SIMDEGA carved out of GUMLA district; **Madhya Pradesh:** In 2003, ANUPPUR district created from SHAHDOL district, In 2013, AGAR carved out of SHAJAPUR; **Nagaland:** In 2004, LONGLENG was carved out of TUENSANG district; **Odisha:** In the year 2000 KHURDA district name was changed to KHORDHA; **Tripura:** In 2012, KHOWAI and SEPAHIJALA was carved out of WEST TRIPURA, GOMATI carved out from SOUTH TRIPURA; **West Bengal:** In 2002, PURBA & PASCHIM MEDINIPUR was formed from MEDINIPUR district.

The distribution of *Pf*DHFR mutations prevalence (Table S1b), its forest plot (Figure 3a) and prevalence trend, where available (Figure 3b), show that all geographical sites are not covered uniformly across the study period. The two *Pf*DHFR mutations, C59R (62%) and S108N (74%), are the most prevalent mutations (pooled estimates 61% and 71%, respectively) and appear to be stabilized/fixed, particularly over the years 2010-17. Along with N51I, these are also the most studied mutations and I164L, the least studied and the least prevalent (8%; pooled 6%). Although rarest overall, the prevalence of I164L was observed to be as high as 32% (2008), 28% (2013) and 12% (2014), presumably due to a very high I164L prevalence in AN (70%; 2008); and AR (70%; 2013 and 46%; 2014). The NES continued to report a high I164L prevalence throughout: ME (36%; 2010; n=14), AR (as above), AS (10%; 2014), NA (37%; 2015; n=38), and TR (100%; 2016; n=17). Relatively higher prevalence of I164L in some of the *P. vivax* dominant locales (Delhi >15%; 2008 and 2010; Haryana 13%; 2015) raises concerns as AS+SP is still the treatment of choice for *P. falciparum* malaria in these areas.

**Figure 3a.**
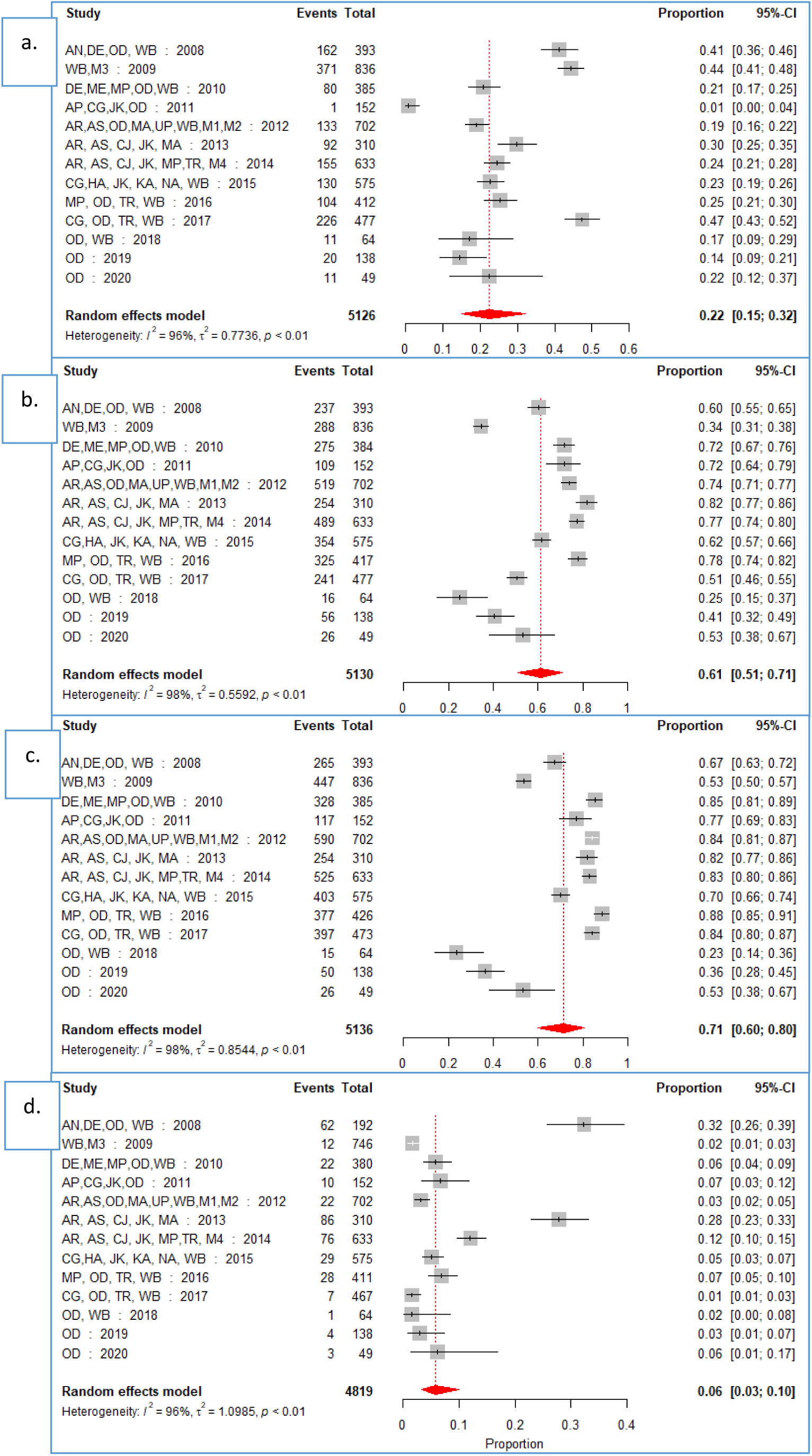
Forest plot of *Pfdhfr* mutations. Pooled prevalence along with 95% CI of individual mutations in four different loci of *Pf*dhfr gene (a: N51I; b: C59R; c: S108N; d: I164L) is shown in different States/UTs and arranged in increasing chronological order of data collection years from 2008 to 2020. Here, “Events” and “Total” represent the number of number of samples with specified mutation and number of samples screened for mutations, respectively. The data are arranged in ascending order of start year of data-collection. AN: Andaman and Nicobar Islands; AP: Andhra Pradesh; AR: Arunachal Pradesh; AS: Assam; CG: Chhattisgarh; DN: Dadra and Nagar Haveli; GU: Gujarat; GO: Goa; DE: Delhi; HA: Haryana; JK: Jharkhand; KA: Karnataka; MA: Maharashtra; ME: Meghalaya; MI: Mizoram; MP: Madhya Pradesh; NA: Nagaland; OD: Odisha; PU: Punjab; RA: Rajasthan; TR: Tripura; UP: Uttar Pradesh; WB: West Bengal

Although the burden of *Pf*DHFR C59R and S108N, has been ≥60% throughout barring 2009 & 2018-20, ≥90% isolates had S108N in AN (2008), ME/MP/WB (2010), JK (2011), UP/WB (2012), AR (2013-14), NA/WB (2015), MP/TR/WB (2016) and TR (2017). UP, MP and WB are areas to focus as the prevalence of S108N was found >60% every year. For C59R, the overall annual prevalence was ≥60% from 2008-2016, barring 2009 (34%) 2017 & 2020 (∼50%). More than 70% mutant isolates were reported from AN (2008), ME/MP/WB (2010), JK (2011), WB (2012), AR/AS (2013-14), MP/TR (2014), NA (2015), MP/TR/WB (2016), OD/WB (2017). The overall burden of N51I mutation was 29% (pooled 22%) with >40% prevalence in 2008-09 and 2017 with alarming burden in AN (70%, 2008), WB (69%, 2008; 74%, 2009; 66%, 2015; 50%, 2017), AR (69%, 2013), TR (>50%, 2016-17), and CG (58%, 2017). The *Pf*DHFR mutations prevalence trend (Figure 3b) shows the need for continuous monitoring of SP-resistance markers specially in states other than the NES and specially in OD with rising trends in all 4 loci.

**Figure 3b.**
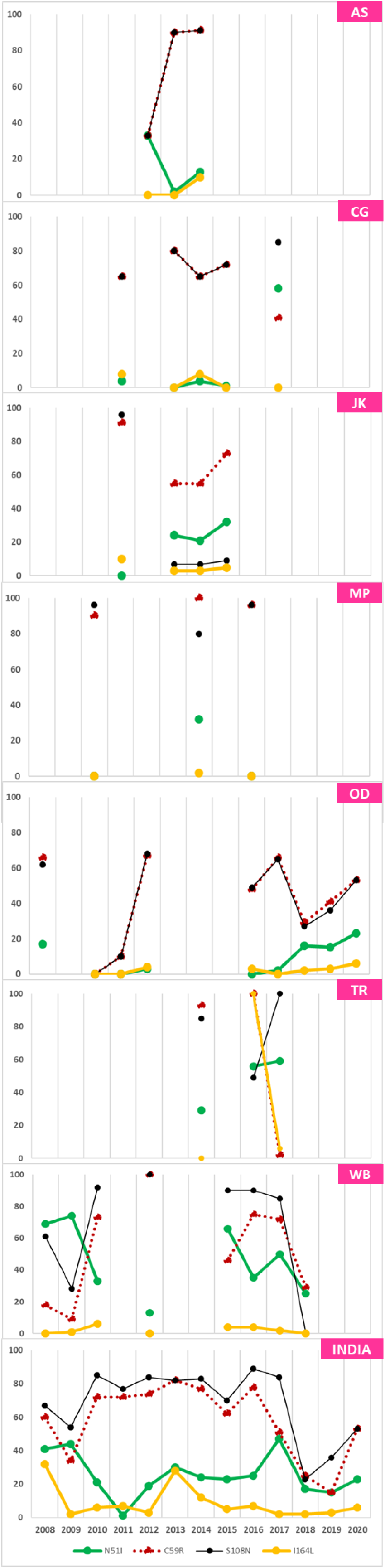
Line diagram showing trends of *Pf*DHFR mutation from 2008-2020. X-axis denotes year of study and Y-axis denotes percentage of mutation in *Pf*DHFR. Distribution of *Pf*DHFR mutation prevalence (%) covered in three or more different years with data breakup are shown alphabetically Assam (AS), Chhattisgarh (CG), Jharkhand (JK), Madhya Pradesh (MP), Odisha (OD), Tripura (TR), West Bengal (WB) and India as a whole, respectively.

The *Pf*DHFR double mutants (Table S1d) were the most prevalent (51%; pooled 42%) overall. Individual locales that showed >70% double mutants include MP (90%) and WB (78%) in 2010, WB (75%) in 2012, AS (90%) in 2013, AR (77%) and AS (73%) in 2014, CG (77%) in 2015, and MP (95%) in 2016 highlighting that MP, WB, and CG are the localities for concern for surveillance as SP is not in use in NES as a part of ACT since 2014. The overall prevalence of triple and quadruple mutations was 6% and 5%, respectively which is also an area of immediate concern for states like WB (8%, 2008; 12%, 2012), OD (12%, 2019; 8%, 2020) and CG (16%, 2017) for triple mutants and AN (68%, 2008), WB (2% in 2017) and OD (2%, 2017 & 2020) for quadruple mutants.

For *Pf*DHPS mutations (Table S1c, Figure 4a-b), the most prevalent mutation reported was A437G (39%), followed by K540E (25%) and A581G (12%) but the pooled estimates were 32%, 22% and 12%, respectively. However, the state-wise breakup for prevalence of these mutations clearly shows that some states have much higher prevalence of these mutations: WB (A437G/K540E); MA (A437G; 2012-13); AR (A437G/K540E/A581G; 2013-15); AS (A437G; 2013, 2015); TR (A437G/K540E/A581G; 2015-17); JK (K540E; 2013, 2015); OD (K540E/A581G; 2017-18, 2020) and DE (K540E; 2008, 2010). The trend shows that the most common *Pf*DHPS mutation is A437G with rising and near-100% prevalence in WB (2010-12), and TR (2015-17). All 3 mutations show a rising trend in OD with rising trends from 2016-2020. However, the data for assessing the state-wise trends of the three *Pf*DHPS mutations is scattered and incomplete.

**Figure 4a.**
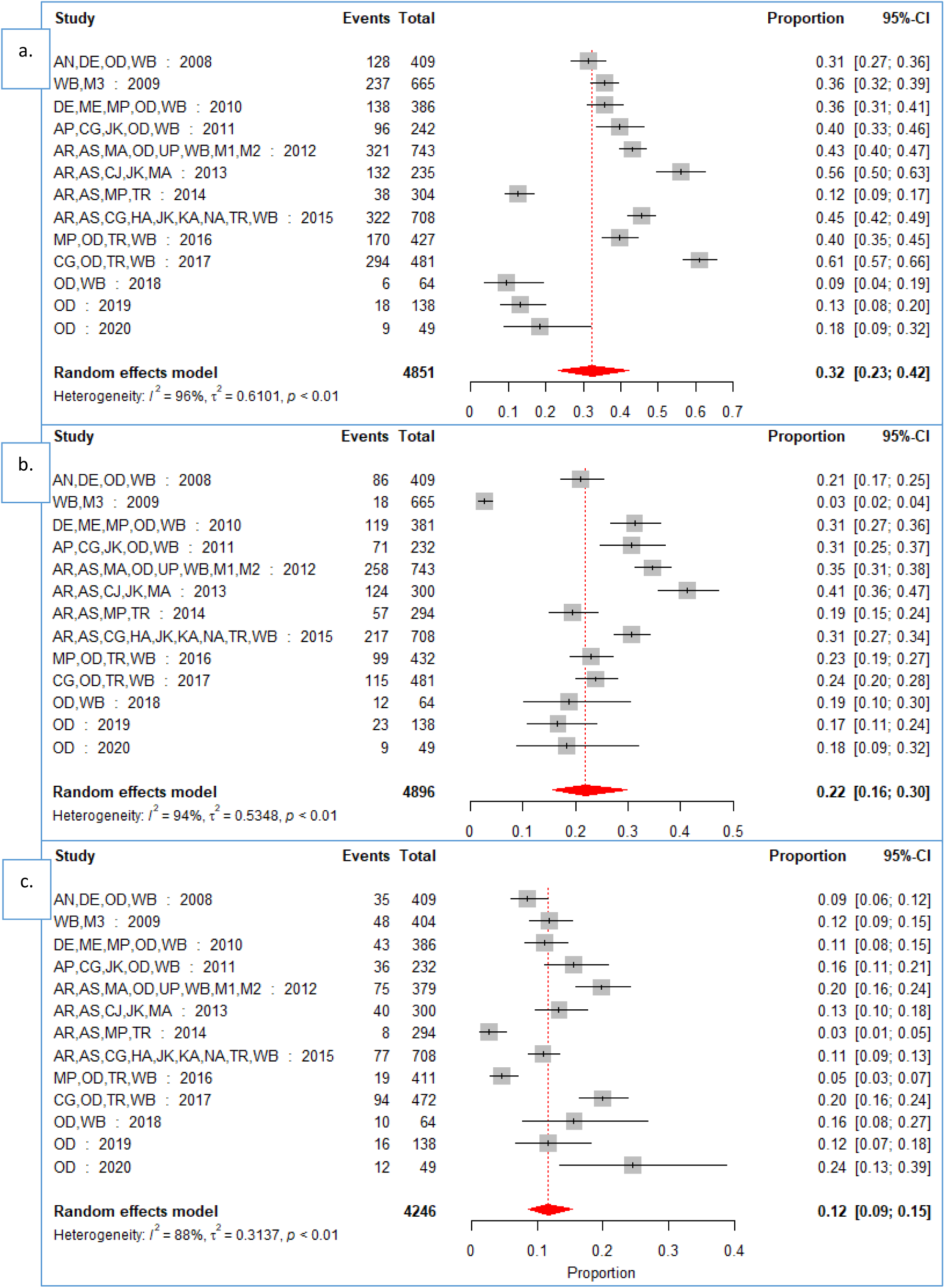
Forest plot of Pfdhps mutations. Pooled prevalence along with 95% CI of individual mutations in three different loci of Pfdhps gene (a: A437G; b: K540E; c: A581G) is shown in different States/UTs and arranged in increasing chronological order of data collection years from 2008 to 2020. Here, “Events” and “Total” represent the number of number of samples with specified mutation and number of samples screened for mutations, respectively. The data are arranged in ascending order of start year of data-collection. AN: Andaman and Nicobar Islands; AP: Andhra Pradesh; AR: Arunachal Pradesh; AS: Assam; CG: Chhattisgarh; DN: Dadra and Nagar Haveli; GU: Gujarat; GO: Goa; DE: Delhi; HA: Haryana; JK: Jharkhand; KA: Karnataka; MA: Maharashtra; ME: Meghalaya; MI: Mizoram; MP: Madhya Pradesh; NA: Nagaland; OD: Odisha; PU: Punjab; RA: Rajasthan; TR: Tripura; UP: Uttar Pradesh; WB: West Bengal

**Figure 4b.**
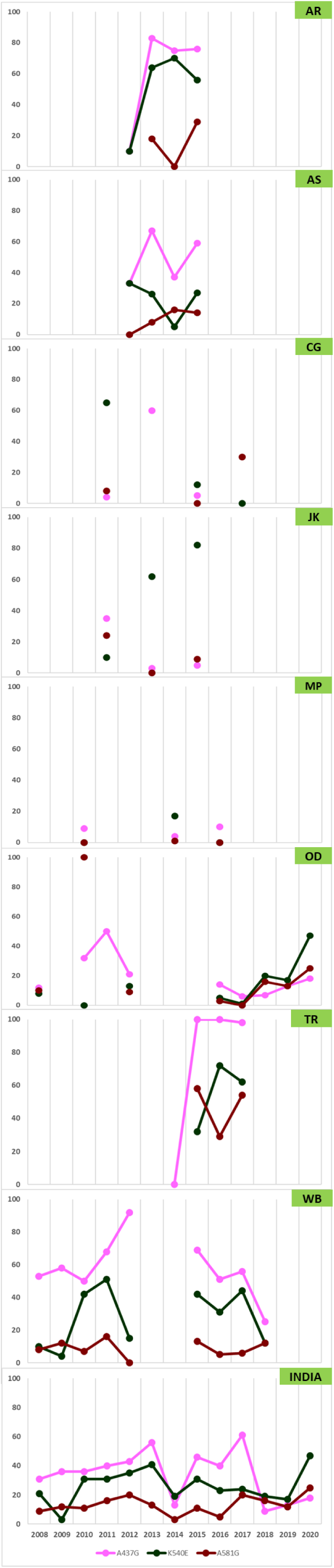
Line diagram showing trends of *Pf*DHPS mutation from 2008-2020. X-axis denotes year of study and Y-axis denotes percentage of mutation in *Pf*DHPS. Distribution of *Pf*DHPS mutation prevalence (%) covered in three or more different years with data breakup are shown alphabetically Arunachal Pradesh (AR), Assam (AS), Chhattisgarh (CG), Jharkhand (JK), Madhya Pradesh (MP), Odisha (OD), Tripura (TR), West Bengal (WB) and India as a whole, respectively.

For *Pf*DHFR/*Pf*DHPS quintuple and sextuple mutations (Table S1d), it was observed that despite a low overall prevalence of these mutations (8% and 0.3%, respectively), states like WB (quintuple >6% in 2008/2010/2012 and >25% in 2015-17), and AN (sextuple 5%; 2008) are worthwhile to consider for continuous surveillance. Not many studies focus on studying quadruple, quintuple and sextuple mutations as the sample size dropped by ∼11% for quintuple/sextuple mutations versus that for triple mutations (Table S1d). The pooled estimates (Figure 5a) of prevalence of single, double, triple and quadruple *Pf*DHFR mutations were 6%, 42%, 4% and 2%, respectively which suggest that the *Pf*DHFR double mutations might be approaching population-level fixation with 75% prevalence in ME, MP, OD and WB in 2010 and higher than average prevalence in 2012-17. The quintuple/sextuple pooled prevalence estimates were relatively much lower, between 0% (sextuple) and 4% (quintuple), but isolated instances of higher quintuple prevalence were observed in 2017 (18%), 2016 (17%), 2015 and 2010 (10%) and 2012 (8%). For sextuple mutations, the years 2017 (CG/OD/TR/WB) and 2008 (AN/DE/OD/WB) reported 1% prevalence each. The prevalence of quintuple mutations, specially from 2015 to 2017 and that of sextuple mutations in 2017 is noteworthy as quintuple/sextuple mutations indicate a very high level of SP-resistance. The trend analyses (Figure 5b) also clearly shows the consistent higher magnitude of double mutations, particularly in 2010 that appears to be contributed by WB which also showed similar high-burden trends in 2017 and 2018. AS (2013), CG (2013/2015/2017), MP (2010/2016), OD (2012-13) and WB (2010/2012/2018) showed a very high double-mutation burden. State-wise trends of other mutations cannot be comprehended as continuous availability of data across the years was unavailable and is an important bottleneck in correct interpretation of mutation trends. The trends, however, indicate a dire need for SP-resistance markers surveillance in WB and OD.

**Figure 5a.**
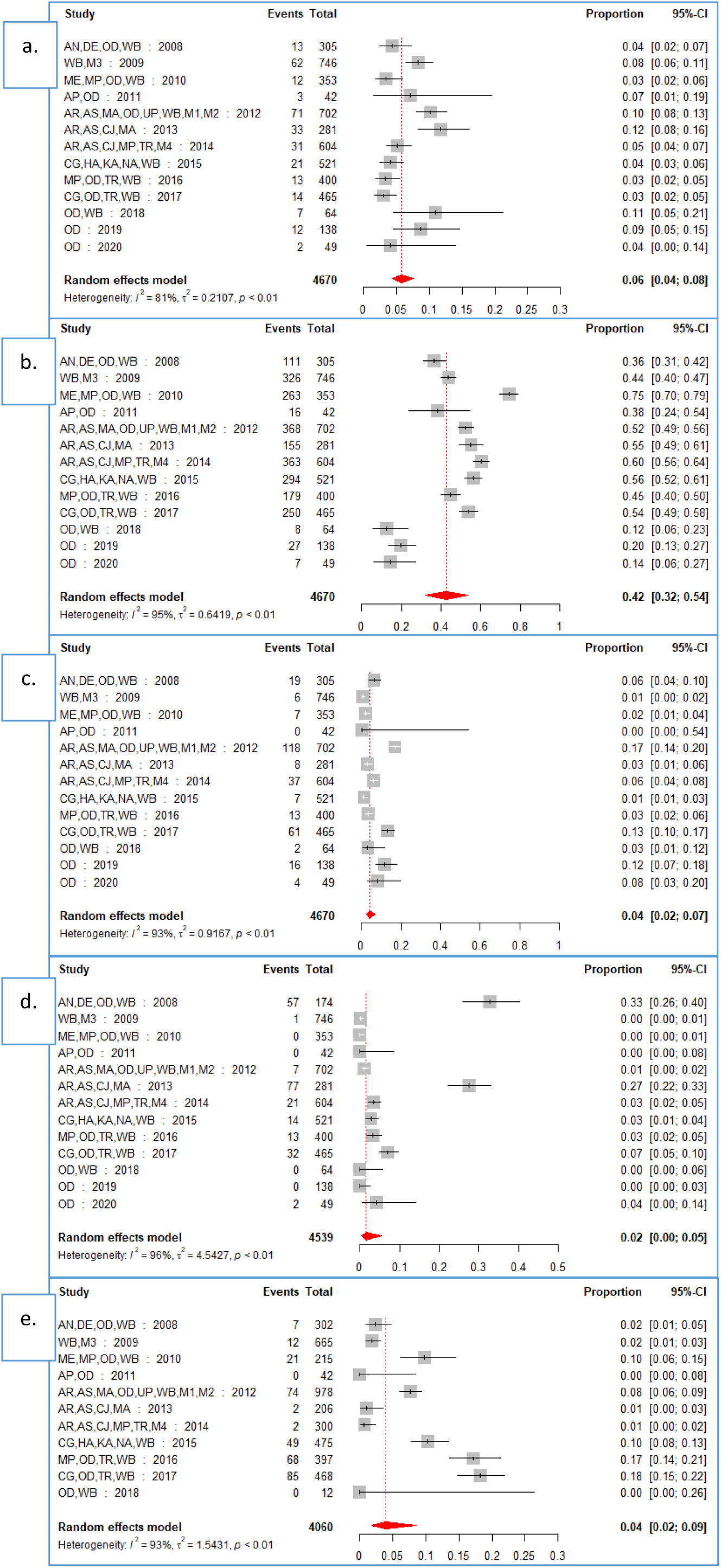
Forest plot of Pfdhfr + Pfdhps mutations taken together. Pooled prevalence along with 95% CI of individual mutations in six different combinatorial loci of Pfdhfr and Pfdhps genes as per standard WHO classification (a: Single (Pfdhfr S108N); b: Double (Pfdhfr S108N + N51I or C59R); c: Triple (Pfdhfr S108N + N51I + C59R); d: Quadruple (Pfdhfr S108N + N51I + C59R + I164L); e: Quintuple (Pfdhfr Triple mutations + Pfdhps A437G and K540E) is shown in different States/UTs and arranged in increasing chronological order of data collection years from 2008 to 2020. Pooled estimates for sextuple (Quintuple + Pfdhps A581G) is not shown due to very low frequencies. Here, “Events” and “Total” represent the number of number of samples with specified mutation and number of samples screened for mutations, respectively. The data are arranged in ascending order of start year of data-collection. AN: Andaman and Nicobar Islands; AP: Andhra Pradesh; AR: Arunachal Pradesh; AS: Assam; CG: Chhattisgarh; DN: Dadra and Nagar Haveli; GU: Gujarat; GO: Goa; DE: Delhi; HA: Haryana; JK: Jharkhand; KA: Karnataka; MA: Maharashtra; ME: Meghalaya; MI: Mizoram; MP: Madhya Pradesh; NA: Nagaland; OD: Odisha; PU: Punjab; RA: Rajasthan; TR: Tripura; UP: Uttar Pradesh; WB: West Bengal

**Figure 5b.**
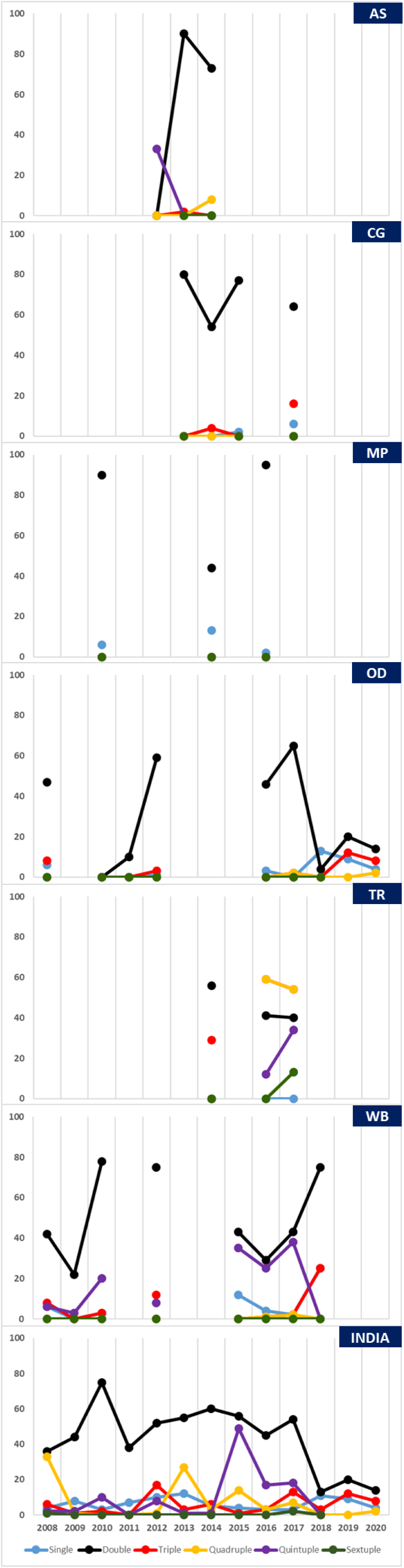
Line diagram showing trends of *Pf*DHFR+*Pf*DHPS mutation from 2008-2020. X-axis denotes year of study and Y-axis denotes percentage of mutation in *Pf*DHFR+*Pf*DHPS. Distribution of *Pf*DHFR+*Pf*DHPS mutation prevalence (%) covered in three or more different years with data breakup are shown alphabetically: Assam (AS), Chhattisgarh (CG), Madhya Pradesh (MP), Odisha (OD), Tripura (TR), West Bengal (WB) and India as a whole, respectively.

India harbours a total of 17 mutant *Pf*DHFR haplotypes (Figure 6a) and NRNI was found to be the most common haplotype across the country, whereas IRNL was found only in the NES and AN. IRNI was found to be concentrated towards the north-eastern and eastern parts of India (AS/AR/TR/NA/WB/JK/OD/CG/AN), whereas NCNI was found only CG/HR/MA/MP/OD/JK/UP. NCSL, NRSL and ICII haplotypes were unique to HA, OD and WB, respectively. The maximum number of haplotypes were reported in WB (12/17; 71%) followed by OD (8/17; 47%) and CG (7/17: 41%) and the least in KA (1/17: 6%) which might be indicative of differential drug pressure. On the contrary, 36 *Pf*DHPS mutant haplotypes (Figure 6b) were found. The most common, AGEAA, was found in the entire country except AR/MP/KA/HA. No *Pf*DHPS mutations were reported in UP whereas maximum haplotypes were reported from WB (24/36; 67%) and AR (12/36; 33%) and the least in ME (1/36) probably because of low sample size. The absence of *Pf*DHPS mutations in UP may indicate that the parasite population has not been sampled enough as *Pf*DHFR mutations were reported from UP. The *Pf*DHPS haplotype data is insufficient from RA/MA/GU/DE to make conclusive statements.

**Figure 6a.**
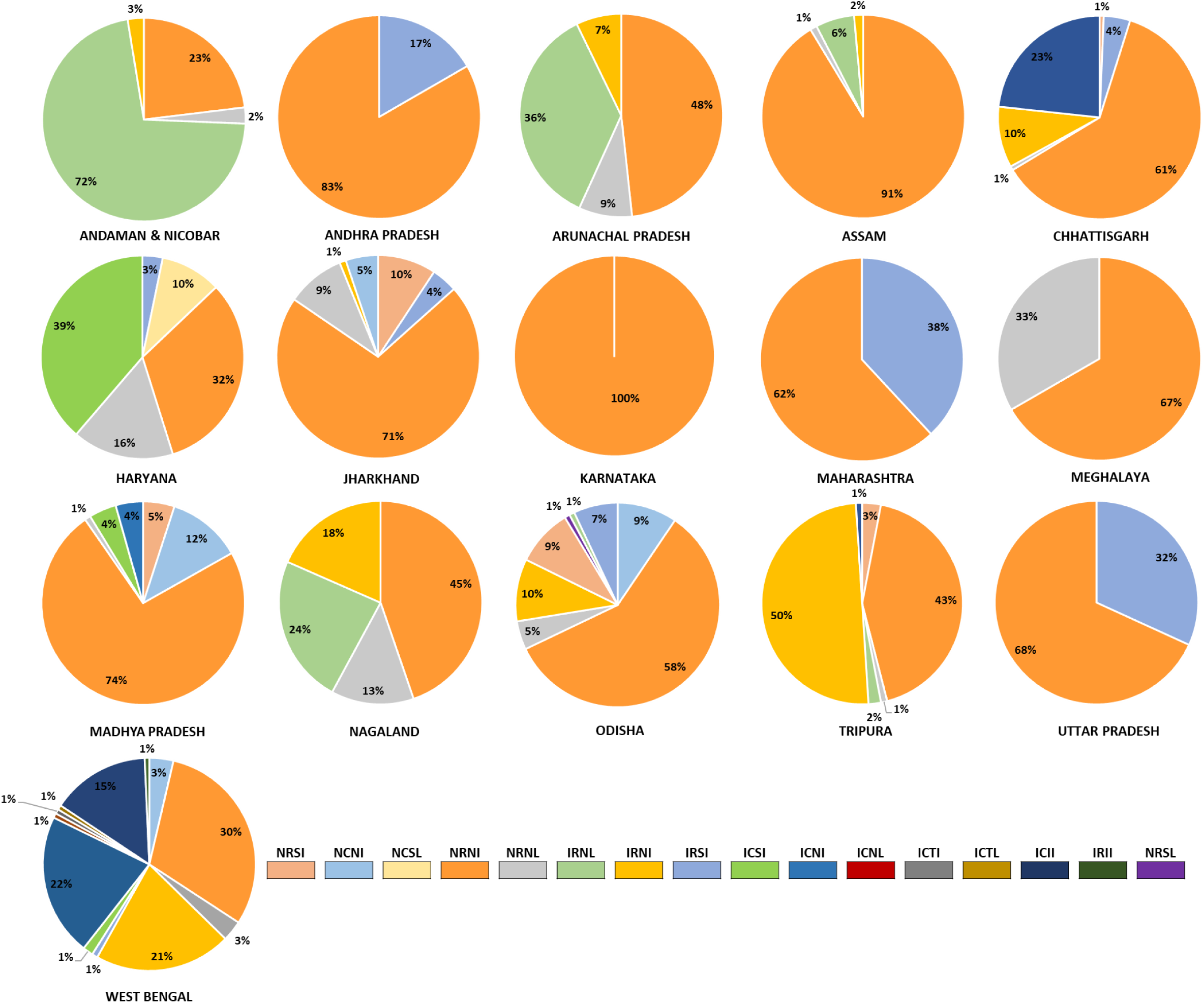
Pie-diagram showing distribution of 16 *Pf*DHFR mutant haplotypes present in different states across India. The states have been arranged in alphabetical order. A total of 17 haplotypes (excluding the wild type, NCSI) were reported across India. This figure excludes NCNL, a haplotype which was reported in a study with no available data break up (details available in supplementary table). NRNI is the most common haplotype across the country, whereas IRNL was present only in the north-eastern parts of India, Odisha and Andaman & Nicobar Islands. West Bengal harboured maximum number of haplotypes (12/17) followed by Odisha (8/17) and Chhattisgarh (7/17) and the least was reported from Karnataka (1/17). NRSL haplotype has recently been reported only in Odisha in 2019. ICII is another haplotype reported from WB only.

**Figure 6b.**
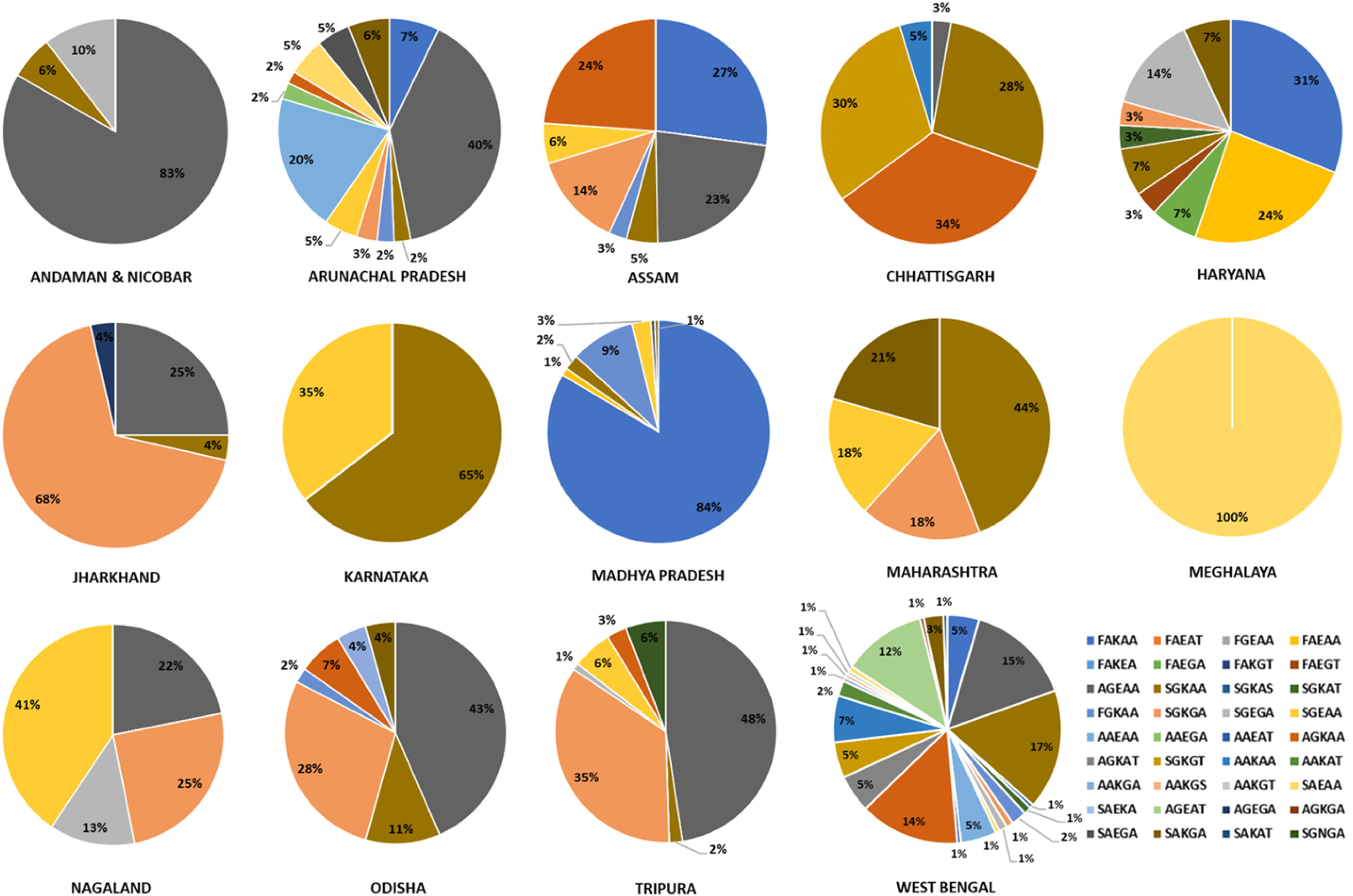
Pie-diagrams showing distribution of 36 *Pf*DHPS haplotypes present in different states across India. The states have been arranged in alphabetical order. A total of 36 haplotypes (excluding the wild type, SAKAA) were reported across India. The most common haplotype was AGEAA found in entire country except in Andhra Pradesh, Madhya Pradesh, Karnataka, and Haryana. No haplotypes have been reported from Andhra Pradesh and Uttar Pradesh whereas the maximum number of haplotypes were reported from West Bengal (24/36) followed by Arunachal Pradesh (12/36). Meghalaya reported the least number of haplotypes (1/36).

## 4. Discussion

The selection and spread of SP-resistance has been faster ^33^ than CQ-resistance ^34^ and hence the need to monitor SP-resistance. With increasing tendency of travel, parasites carrying the high-resistance conferring *Pf*DHFR/*Pf*DHPS mutations can be easily translocated. This could be particularly dangerous if such translocation happens along the SP-usage differential from high-prevalence foci (SP not currently used) to areas with low prevalence of such mutations but with ongoing SP drug pressure thus favouring their faster selection and transmission.

Although a drug policy change is recommended with >10% treatment failures ^4, 18^, isolated monitoring of SP-resistance markers ^4^ provide the earliest evidence of emerging drug resistance and therefore offer a chance for primordial prevention of establishment of resistance. ^4, 25, 35^ This is even more important in regions where TES cannot be regularly conducted ^3, 36^ and/or has additional challenges ^37, 38^.

No comprehensive systematic review and meta-analysis on SP-resistance markers prevalence has been reported making this study, the only such study in the region to date. The study also gains paramount importance in light of the flags raised towards emerging ‘artemisinin resistance’ in eastern India. ^39–42^

To date, we could find only three published reports ^26–28^ that tried to summarise the geo temporal trends of SP-resistance and/or their markers in India. Shah et al. systematically reviewed *P. falciparum* SP-resistance from 1997-2007 through TES but avoided molecular data. Arya et al. did a systematic review that included clinical and molecular data on ACT resistance in India and Africa published between 2008 and 2018 whereas Chaturvedi et al. published a narrative review on the structural basis of SP-resistance conferring mutations in *P. falciparum* and *P. vivax* and their global spatio-temporal trends between 2005 and 2019, including that in India.

The current report is unique as it not only covers the data from studies included by Arya et al. and Chaturvedi et al. but has certain striking positive differences. The current study is a meta analysis of published/unpublished data (from 2008 to January 2023) on *Pf*DHFR/*Pf*DHPS resistance-conferring mutations as reported from India. The analysis grants comparability in time and space as WHO-recommended standardised definitions of mutation classifications were considered and data were categorised, reorganized and reclassified accordingly. Whereas Arya et al., just provided a qualitative statement of the number and type of studies that evaluated SP-related mutations, they did not analyse/present the data using quantitative summary measures. The present study also has avoided ambiguity and incomparability that were observed in published literatures ^3, 28^ and online SP-molecular surveyor ^43^.

The classification of SP mutations from single to sextuple is sometimes done based simply on the numerical count and combinations of any independent mutations in *Pf*DHFR/*Pf*DHPS. This is in contrast to the WHO’s recommendation to use specific *Pf*DHFR/*Pf*DHPS mutations to classify them as single to sextuple mutations. This has been observed in many recent publications ^3, 28^ where the comparability of mutation data between different states of India has not only been compromised but has also incorporated errors in interpretation. ^3^ Further, the term “prevalence” of mutations appears to be slightly misrepresented by Chaturvedi et al. wherein they calculated the proportion of specific SP-mutations out of the total observed mutations which might be misleading when it comes to determining prevalence in any epidemiological studies. The present meta-analysis also avoids this ambiguity and chances of misinterpretation of terminology.

Chaturvedi et al. includes proportion data on SP-resistance mutations from 2005-2019 but due to lack of data-breakups, they were unable to expand the data coverage across India. This lacuna has also been filled by the present study by rigorous communication with concerned authors and collecting data-breakups from 2008 onwards.

Since Chaturvedi et al. and the current study use two different endpoint measures, proportion and prevalence, respectively, the data on single, double and triple mutations cannot be compared. However, the important areas representing critical burden of SP-mutations do overlap as both studies show that the burden of quadruple mutants is driven by AN (∼87% proportion; ∼68% prevalence), and AR (∼37% proportion; ∼36% prevalence). Both the studies addressed quintuple and sextuple mutations following comparable definitions. But, while the current study shows the presence of quintuple mutations in AN (2008), AR (2012-14) and WB (2008-10/2012/2015-17), Chaturvedi et al. reported quintuple mutation only in AR (Changlang) in 2013. Chaturvedi et al. did not report any sextuple mutations in main text or supplementary files but depicted sextuple mutations in one of the figures in AS (Hailakandi and North Lakhimpur). Our study found sextuple mutations in AN (2008) and more recently in TR (2017). Rahi et al. also reported sextuple mutations in AS (2014) and WB (2014-16) but the definition used for sextuple mutation was ambiguous and supplementary data did not correlate with the sites shown in the figure, except for AS and WB. ^3^

Despite some intrinsic limitations including non-availability of state-wise breakup for studies M1 (2012; n=292); M2 (2012; n=155); and M3 (2009; n=342), this study brings forth certain challenges and suggests the way ahead (Box 1).

## 5. Conclusion

With delayed parasite clearance by artemisinin, high SP efficacy (and low burden of SP resistance markers) is essential to prevent AS+SP therapeutic failures. This exhaustive spatio temporal meta-analyses highlights the need for SP-resistance markers’ surveillance in India and concludes that the following geographical areas and mutants warrant prioritized molecular surveillance for *Pf*DHFR/*Pf*DHPS singular/combinatorial mutations: UP/MP/WB (single); MP/WB/CG (double); AN/WB/OD/CG (triple/quadruple); WB/AN (quintuple); and AN/CG/OD/WB/DE (sextuple). This study classifies these areas into hot spots where such surveillance would be most crucial (Figure 7). These include districts classified into 5 clusters where a continuous vigil is warranted: AN, AP, AR, AS, CG, HA, MP, NA, OD, TR, UP and WB. Evidence from such malaria genetic surveillance ^44–48^ may be used as a reference model to develop a framework in India. Countries aiming malaria elimination have been strongly advised to adopt genetic surveillance as an intervention. ^49, 50^

**Figure 7.**
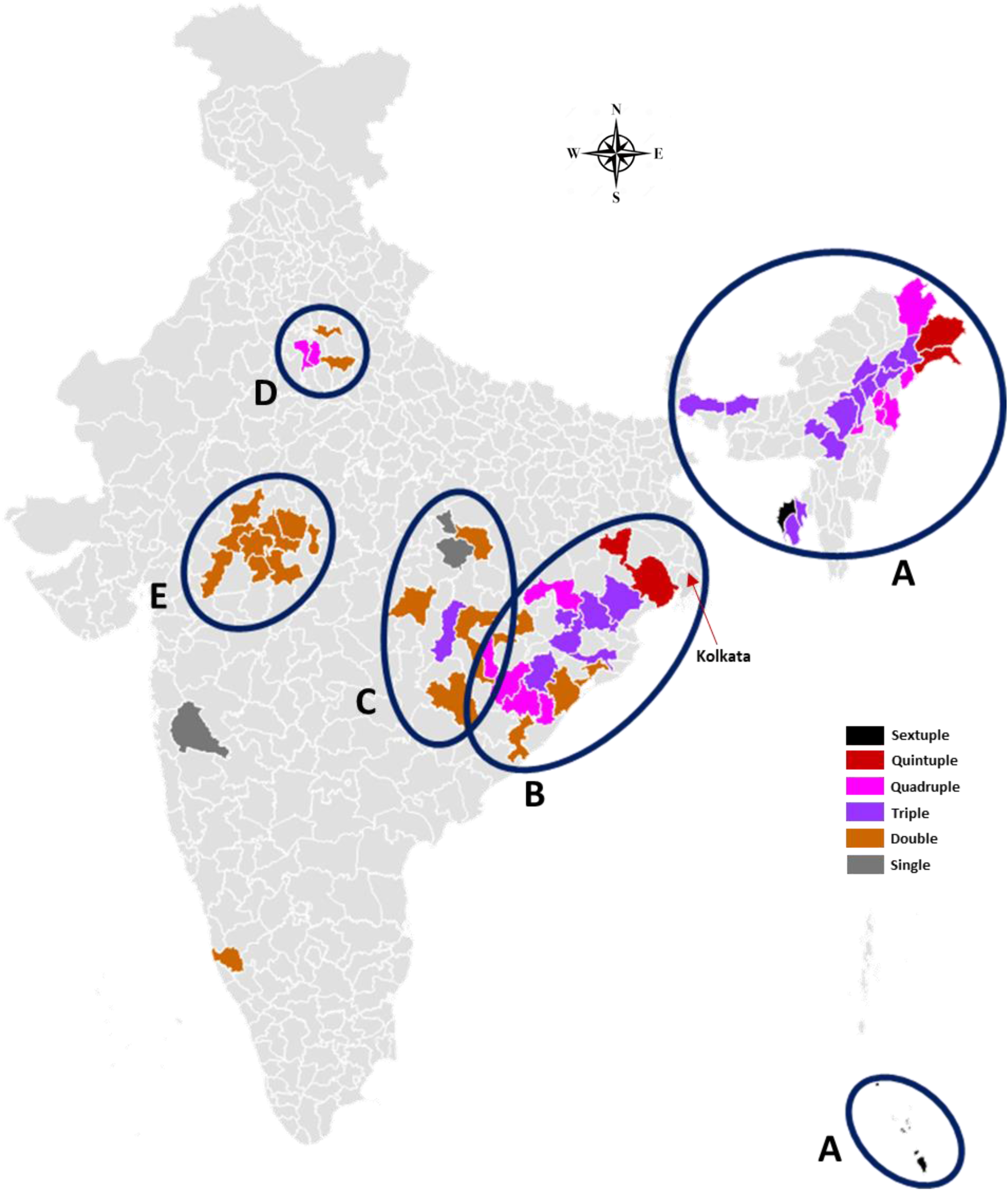
District map of India showing SP-resistance markers hotspots (created with https://gramener.com/map/). Data encapsulated here is based on the prevalence of *Pfdhfr*+*Pfdhps* WHO-validated SP-resistance markers across the country (Table 5). The criteria for classifying a district as a hotspot for a particular mutation is prevalence of the mutation ≥ lower bound of the country’s pooled estimate’s 95% CI as obtained from Figure 6a. Hence, the threshold is the prevalence ≥2% for single, ≥46% for double, ≥2% for triple, ≥1% for quadruple, and >0% for quintuple and sextuple mutations. The hotspot-districts are organized into 5 clusters (A-E, from east to west) based on the presence of the mutations conferring the highest order of resistance. ***Cluster A (sextuple and below):*** Sextuple - TR (West Tripura); AN (Nicobar); Quintuple - AR (Changlang, Lohit); Quadruple - AR (Lower Dibang Valley, Tirap); NA (Dimapur, Longleng, Mokokchung, Zunheboto); Triple - TR (Dhalai, Khowai, West Tripura, South Tripura); AS (Chirang, Dibrugarh, Golaghat, Jorhat, Karbi Anglong, Lakhimpur, NC Hills, Sivasagar, Tinsukia); WB (Jalpaiguri). ***Cluster B (quintuple and below):*** Quintuple - WB (Kolkata, Medinipur, Purulia); Quadruple - OD (Ganjam, Kalahandi, Kandhamal, Nuapada, Rayagada, Sundergarh); Triple - OD (Angul, Cuttack, Debagarh, Keonjhar, Mayurbhanj); Double - AP (Vizianagram); OD (Bargarh, Ganjam, Khordha). ***Cluster C (triple and below):*** Triple - CH (Durg); Double - CH (Bastar, Koriya, Raipur), MP (Balaghat); Single - MP (Anuppur). ***Cluster D (quadruple and below):*** Quadruple - HA (Mewat); Double - UP (Aligarh, Ghaziabad). ***Cluster E (double):*** MP (Agar, Bhopal, Dewas, Indore, Jhabua, Mandsaur, Rajgad, Ratlam, Ujjain).

The key question is whether the time has come for yet another anti-malaria treatment policy change from AS+SP to AL in the rest of India? The spatio-temporal pattern of validated *Pf*DHFR/*Pf*DHPS mutations that mediate SP-resistance suggests that expansion of AL throughout India as first line therapy for uncomplicated Pf malaria might be a better option as a primordial prevention to widespread fixation of highest level of SP-resistance. On the other hand, the AS+SP therapeutic failure rates do not support this change which is also backed up by evidence suggesting presence of certain compensatory genetic changes (including copy-number variations and selective sweep) in the parasites that are able to salvage the folate pathway despite the drug pressure thus sustaining some of the high SP resistance driving mutations. ^52–54^ The evidence for the risks needs to be weighed but for a country envisaging malaria elimination by 2030, taking the risk of continuing AS+SP, particularly with rising flags of artemisinin-resistance might be counterproductive and the decision needs to be made sooner rather than later.

## Data Availability

All data produced in the present work are contained in the manuscript

## Author Contributions

AS: Conceptually designing the work. Acquisition, analysis and interpretation of data. Critically drafting, editing, revising and finalizing the MS along with figures.

SK: Acquisition, analysis and interpretation of data. Editing and revising the MS critically. Drafting and finalizing the figures and tables of PfDHFR, editing, revising and finalizing the MS along with figures.

CC: Acquisition, analysis and interpretation of data. Editing and revising the MS critically. Drafting and finalizing the figures and tables of PfDHPS.

CP: Statistical data analysis and interpretation. Editing, revising and finalizing the MS.

LK: Acquisition, analysis and interpretation of data. Critically drafting, editing, revising and finalizing the MS.

## Acknowledgement

The authors apologize to colleagues whose work could not be cited owing to the broad scope of the review and space limitation. The authors would like to acknowledge ICMR-NIMR for providing working space in the institute. The authors would particularly like to thank Dr Praveen K Bharti (ICMR-National Institute of Malaria Research, New Delhi), Dr Siraj A Khan (ICMR-Regional Medical Research Center, NE region-Dibrugarh, Assam), Dr Rakesh Sehgal (Post Graduate Institute of Medical Education and Research, Chandigarh), Dr Manuj K Das (ICMR-Regional Medical Research Center, NE region-Dibrugarh, Assam), Dr Jitendra Sharma (District Epidemiologist, Government of India, Lakhimpur, Assam), Dr Amit Kumar (ICMR-National Institute of Malaria Research, New Delhi), Dr Krishanpal Karmodiya (Indian Institute of Science Education and Research, Pune) and Dr Manoranjan Ranjit (ICMR-Regional Medical Research Centre, Bhubaneswar) for their support in providing the details of the data without which a uniform and standardized compilation would not have been possible. LK was funded by US NIAID MESA-ICEMR Program Project. SK and CC were supported by project senior research fellowship from ICMR.

## Data sharing

All data are available as supplementary information.

## Box 1 Challenges and future path

- Standardization of reporting for publishing information on SP-resistance markers

There are certain issues that need to be addressed at the level of researchers. These include publication of data in a uniform WHO-recommended format for reporting PfDHFR/PfDHPS mutations. Issues like non-ambiguous classification of data collection year and month, and mentioning the exact site of data collection need to be addressed at the time of submitting the data for publication. These challenges could be circumvented if a standardized local guidelines are made for reporting information on SP-resistance markers.

- Delayed publication

An average ‘publication delay’ (time from data collection to publication) of 3.5 years (range 1 year to 12 years), as noted in this study (Supplementary table S2), might significantly affect the decision making by the National Control Programs^17^. This is further jeopardized by missing an opportunity to publish at all as a significant chunk of data remains unpublished. It is recommended that such platforms should be developed wherein confidential sharing of such unpublished raw data may be invited in real-time from researchers and evidence can be synthesized to a meaningful and actionable information.

- Genetic surveillance for drug resistance markers

SP therapeutic efficacy may not be monitored alone and hence a robustly designed genetic surveillance of drug resistance markers is recommended.^35^ Isolated monitoring of SP resistance markers may be adopted as TES have their own inherent limitations including meeting the desired sample size with declining malaria cases, operational and logistic challenges of patient screening, recruitment and hospitalization, and treatment failures stemming from certain patient-centric attributes including pharmacokinetic and pharmacodynamics variations and immunity.^55^ In addition, an independent threshold for prevalence of high SP-resistance conferring mutations^18^ should be developed to guide the policy much before the actual widespread appearance of treatment failure.^56^

